# RedFuMOS: A novel approach for multi*-omics* and clinical data-driven patient stratification

**DOI:** 10.64898/2026.08.26.26361415

**Authors:** Sara De Luca, Carmen Fava, Giuseppe Rizzo, Alessia Visconti, Paola Berchialla

## Abstract

**Background:** Patient stratification from multi*-omics* and clinical data is essential for uncovering disease heterogeneity and moving toward more personalized treatment strategies. However, integrating heterogeneous data layers while identifying robust patient strata remains challenging.

**Methods:** We introduce Reduced Fusion of Multi-*Omics* Stratification (RedFuMOS), a novel three-step approach for patient stratification based on mixed-type multi*-omics* data. RedFuMOS extends Similarity Network Fusion to accommodate mixed-type data layers and layer-specific similarity measures for data integration, includes a dimensionality reduction step to mitigate the curse of dimensionality, and performs patient stratification using density-based hierarchical clustering with HDBSCAN. It also implemented an automated optimization procedure to identify the best set of hyperparameters, minimizing the need for manual tuning.

**Results:** RedFuMOS outperformed six state-of-the-art tools for multi*-omics* patient stratification in a comprehensive simulated benchmarking study, which also confirmed that, although computationally expensive, the dimensionality reduction step is crucial for achieving good stratification performance. Additionally, RedFuMOS identified two clinically relevant patient strata in a small real-world cohort of patients with Philadelphia chromosome-positive chronic myeloid leukaemia.

**Conclusion:** RedFuMOS provides a flexible framework for integrating heterogeneous multi*-omics* and clinical data. RedFuMOS is available as an R package at http://github.com/delucasara/RedFuMOS.

## Introduction

Patient stratification plays a crucial role in multiple biomedical applications, including cancer subtyping [1], as distinct patient strata often reflect underlying molecular heterogeneity, and can inform personalized treatment strategies. [2]. In this context, the integration of multi*-omics* data offers a more comprehensive and nuanced understanding of disease biology than single *-omics* studies [3]. By jointly considering multiple molecular data types, integrative approaches can reinforce common signals across related *-omics* layers (*e.g.*, transcriptomics and proteomics), and combine complementary information from layers capturing different biological processes (*e.g.*, glycomics and metagenomics), improving both the robustness and the biological and clinical relevance of the stratification results.

Despite significant advances in this field [4], existing approaches still exhibit several shortcomings. One major difficulty is the so-called curse of dimensionality, where patient cohorts are typically small relative to the large number of measured features [5]. This high-dimensional, low sample size structure can produce noisy similarity estimates, and unstable cluster assignments. Approaches such as integrative clustering (iCluster) [6] and its extension iCluster+ [7], which combine multiple genomics data type into a common probabilistic model before clustering, may be particularly susceptible to this challenge. Perturbation-based methods such as PINSPlus [8], which performs consensus k-means clustering *via* permutation, improve clustering robustness but do not directly construct a reduced representation of the original feature space.

A further limitation is that most approaches assume linear relationships, limiting their ability to capture more complex, potentially non-linear biological interactions. For instance, integrative non-negative matrix factorization (IntNMF) [9] and MoCluster [10] successfully reduce dimensionality before clustering, but are limited by the assumption of linear representation.

Determining the number of strata represents an additional challenge. Some methods, including IntNMF, and MoCluster, require the number of strata to be known in advance, although this information is often unavailable. A potential solution is to incorporate data-driven procedures for selecting the number of strata. For example, the Cancer Integration via Multi-kernel LeaRning (CIMLR) approach [11], learns a low-dimensional non-linear representation of the data and includes an optimisation step to select the number of strata.

Missing data constitute a further challenge. Similarity Network Fusion (SNF) [12] addresses missingness by performing k-nearest neighbours imputation within individual data layers. Notably, SNF makes no assumption regarding the underlying data distribution and is agnostic to the number of strata. However, it does not perform data reduction, and may remain sensitive to noisy or high-dimensional input data.

Finally, most established multi*-omics* stratification approaches are designed for numerical molecular data, and cannot accommodate mixed-type clinical layers without additional preprocessing. This limits the direct integration of clinical characteristics, including binary, ordinal, and categorical variables such as sex, ancestry, treatment response, and gene mutation, which are often critical for patient stratification [13].

To overcome these limitations, we introduce Reduced Fusion of Multi-*Omics* Stratification (RedFuMOS), a novel three-step approach for mixed-type multi*-omics* patient stratification (**Fig. 1**). RedFuMOS is freely available as an open-source R package at http://github.com/delucasara/RedFuMOS.

**Fig. 1.**
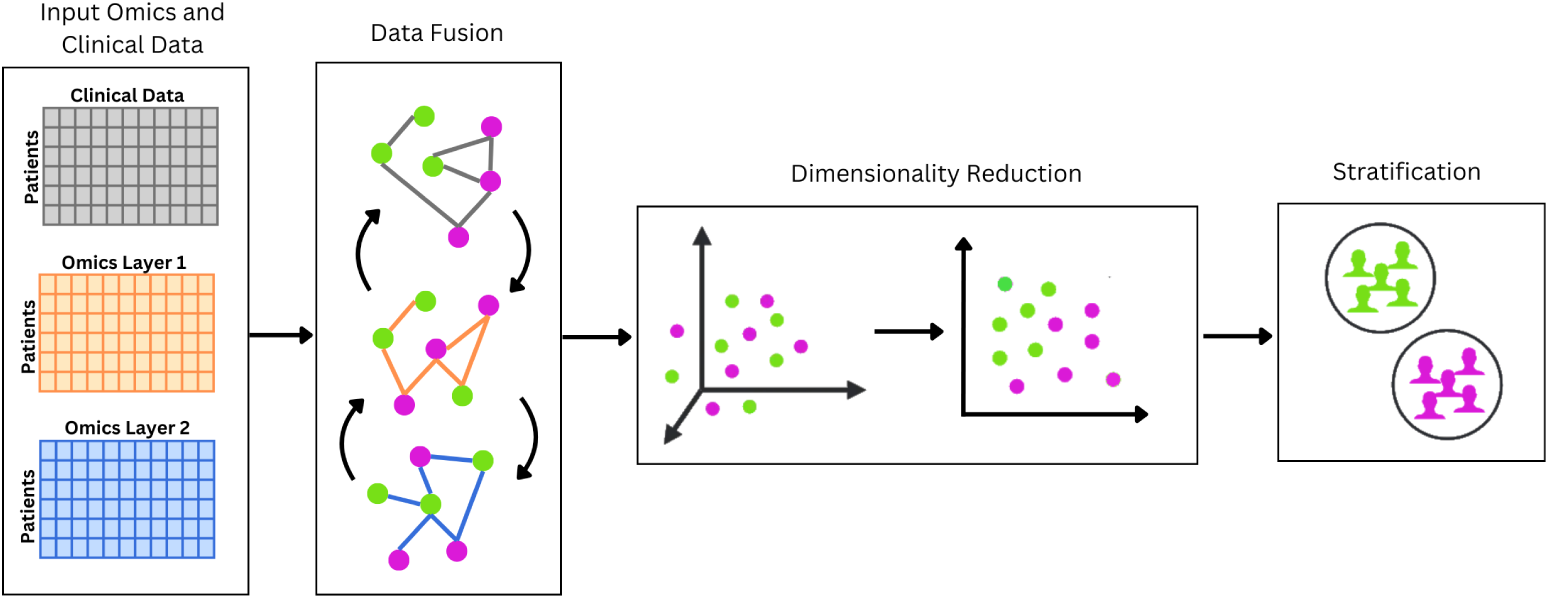
RedFuMOS workflow. Using multiple *-omics* layers as well as mixed-type clinical data as input, RedFuMOS first performs data fusion using SNF. It then applies dimensionality reduction to the fused data matrix using either PCA, UMAP, or t-SNE. Finally, it performs patient stratification using density-based hierarchical clustering (HDBSCAN). RedFuMOS includes an optimisation procedure that automatically selects the hyperparameters yielding the best internal stratification performance.

## Methods

### RedFuMOS implementation

RedFuMOS consists of three main steps: data fusion, dimensionality reduction, and patient stratification. We use an automated procedure for hyperparameter optimization to select the configuration that provides the best internal stratification performance.

#### Data fusion

To avoid assumptions about data distribution and handle missing data, we integrated multi*-omics* and clinical data layers into a unified network representation using SNF [12] (SNFtool R package; version 2.3.1). Briefly, SNF starts by imputing missing values using the k-nearest neighbours (k-NN) approach. Then, to reduce differences in scale across variables, it normalizes each *-omics* layer. Next, it calculates a patient-by-patient distance matrix for each layer using the Euclidean distance. The resulting distance matrices are then transformed into affinity matrices. Within each affinity matrix, SNF uses a k-NN procedure to identify layer-specific, strongly connected sub-network structures. These subnetworks are then fused into a single patient-by-patient affinity matrix that integrates information across all input layers.

Here, we extended the original SNF algorithm to calculate a patient-by-patient distance matrix using a measure appropriate to each data layer, enabling a more accurate modelling of the different characteristics underlying each *-omics* dataset. By default, we use cosine distance, which has been shown to outperform other measures in similarity network construction [14], while being robust to outliers and sparse data. Then, following recommendations from the literature, we implemented the Gower distance [15] for clinical information, which often includes mixed-type data, and which could not be directly used in the original SNF implementation; the Bray-Curtis dissimilarity for microbiome data [16]; the Euclidean distance for DNA methylation [17]; and the Pearson correlation for gene expression [18], protein levels [19], and metabolomic data [20]. Pearson correlations were converted to distances using the formula: distance = 1 *−* correlation. We set the self-similarity (main diagonal) to zero.

#### Dimensionality reduction

To obtain a lower-dimensional representation of the fused patient-similarity network, we implemented three dimensionality reduction approaches: principal component analysis [21] (PCA; function prcomp, stats R package, version 4.4.2), t-distributed Stochastic Neighbor Embedding [22, 23] (t-SNE; Rtsne R package, version 0.17), and Uniform Manifold Approximation and Projection [24] (UMAP; umap R package, version 0.2.10). PCA is a linear reduction approach that identifies a set of orthogonal features (principal components) while preserving the maximum amount of variance in the original data. By contrast, t-SNE is a non-linear approach that preserves local neighbourhood relationships in a lower-dimensional space. t-SNE maps patients with similar high-dimensional representations to nearby points in the embedding. UMAP is another non-linear reduction approach that works similarly to t-SNE, while further moving dissimilar observations far away from each other.

#### Patient stratification

We input the resulting patient embeddings into the Hierarchical Density-Based Spatial Clustering of Applications with Noise (HDBSCAN) algorithm [25] (dbscan R package, version 1.2.2). HDBSCAN is a density-based clustering method that does not require the number of strata to be specified beforehand, and it is well-suited to high-dimensional spaces and robust to outliers.

#### Automated hyperparameter optimization

To account for the differences in sample size, data structure, and separation between patients, we implemented an automated optimization procedure to identify the set of hyperparameters maximizing the Silhouette score [26]. Specifically, we performed an exhaustive grid-search over the following hyperparameters: for the k-NN algorithm, the number *k* of nearest neighbours (set to range from 5 to *N/*2, with step 1, where *N* is the sample size); for the dimensionality reduction step, the approach to use (PCA, t-SNE, UMAP); and for HDBSCAN, the distance threshold *ɛ*, used for cluster merging (set to range from 0.1 to 1, with step 0.1), and the minimum cluster size (set to range from 5 to *N −* 1, with step 5). Additionally, for dimensionality reduction approaches with tunable hyperparameters, we performed automated optimization based on the trustworthiness score [27], which measures how well the low-dimensional embedding preserves local neighbourhood structure. Specifically, for t-SNE, we optimized the perplexity parameter (set to range from 5 to the smallest value between 50 and (*N −* 1)*/*3, with step 5), and for UMAP the n neighbors parameter (set to range from 5 to the smallest value between 50 and *N −* 1, with step 5) and the min dist parameter (set to take the following values: 0.1, 0.2, 0.3, 0.4, 0.5).

### *In silico* validation

First, we benchmarked RedFuMOS against six state-of-the-art approaches, *i.e.*, iCluster+ [7], PINSPlus [8], IntNMF [9], moCluster [10], CIMLR [11], and SNF [12], all of which have publicly available R packages (**Supplementary Table S1**).

We used the InterSIM R package [28] (function InterSIM, version 2.2.0) to generate synthetic datasets which included three *-omics* layers: DNA methylation, mRNA gene expression, and protein expression. InterSIM simulates realistic datasets which preserve the correlation structures observed in The Cancer Genome Atlas (TCGA) ovarian cancer study [29]. Users can simulate datasets with a given number of patients assigned to a given number of strata with given probabilities. Users can also select the degree of separation *δ* between strata.

To perform an exhaustive evaluation during benchmarking, we simulated 20 datasets with different degrees of separation, ranging from 0.1 (poorly separated) to 2 (almost perfectly separated) in steps of 0.1. We fixed the number of patients at 100, and assigned them to four equally sized strata (*i.e.*, we set a 25% probability per stratum). To make the simulation as realistic and fair as possible, we did not provide the number of strata as input. For CIMLR, SNF, and iCluster+ we used the data-driven procedures implemented by each method to determine the number of strata. For all other approaches, we used the same automated hyperparameter optimization procedure based on the Silhouette score implemented in RedFuMOS. To assess the performance stability of the benchmarked methods, we repeated the experiment 30 times using different synthetic datasets, and reported the mean and standard deviation of the resulting Area Under the Receiver Operating Characteristic (AUROC) values.

Next, to assess the scalability of RedFuMOS performance, we generated a series of synthetic datasets including a number of patients ranging from 100 to 1000 in step of 100, divided in four equally sized strata, and across three degrees of separation, namely, 0.1, 0.5 and 1.0. For each dataset, we measured the total running time as well as the running time of each core component: data fusion, dimensionality reduction, and patient stratification. As before, we repeated the experiment 10 times using different synthetic datasets, and reported the mean and standard deviation of the running time.

Finally, we assessed the importance of the computationally expensive dimensionality reduction step by comparing the described version of RedFuMOS with a version in which this step was omitted.

In all experiments, we calculated the similarity matrices using the most appropriate distances, *i.e.*, the Euclidean distance for the DNA methylation layer and the Pearson correlation coefficient for both the mRNA gene expression and the protein expression layers.

All experiments ran on a laptop with Intel i7-1255U 10-core processor and 32GB RAM, using R version 4.4.2.

### Application to real-world data

We evaluated RedFuMOS on a real-world clinical cohort of patients with Philadelphia-positive chronic myeloid leukaemia (Ph+CML) admitted to the Haematology Division at the Mauriziano Umberto I Hospital in Turin, Italy. The cohort included patients receiving active treatment, patients in treatment-free remission, and patients with advanced phases of disease. The study was approved by the Territorial Ethics Committee of *A.O.U. Città della Salute e della Scienza di Torino* (protocol no. CE 209/2022, approval date 2022-05-02). All patients provided written informed consent.

In addition to routine clinical assessments of the efficacy and tolerability of tyrosine kinase inhibitors, we collected demographic characteristics (age, sex) and the plasma levels of trimethylamine N-oxide (TMAO), and of three inflammatory markers, *i.e.*, C-reactive protein (CRP), fibrinogen, and diamine oxidase (DAO). Two *-omics* layers were available. The gut microbiome taxonomic profiling, assessed at the genus level, was measured from 16SrRNA gene amplicon data using the MyMicrobiota’s extraction, sequencing and taxonomical profile protocol (https://www.mymicrobiota.it/). We excluded genera with more than 25% missing data. Peripheral blood cytokine levels were measured with Luminex technology. We excluded cytokines with more than 50% missing data. Patient level distance matrices were calculated using the Bray-Curtis dissimilarity for the microbiome layer, and the Euclidean distance for the cytokines (protein) layer.

We assessed the clinical relevance of the strata identified by RedFuMOS by comparing the proportion of patients achieving a deep molecular response (defined as less than 0.01% BCR-ABL1IS; MR4) across strata using the Fisher exact test. We considered a significance level of 0.05. We further characterized the strata testing for their association with age, sex, and the four previously described plasma biomarkers. We used the Kruskal-Wallis rank sum test for numerical variables and the Fisher exact test for categorical variables. To account for the six comparisons performed, we considered statistical significance using a Bonferroni-adjusted threshold of 0.05/6=0.008.

## Results

### RedFuMOS outperforms competitors across simulated scenarios

In a first experiment, we benchmarked RedFuMOS against six state-of-the-art approaches for multi*-omics*-based patient stratification. To this aim, we simulated 20 scenarios each including 100 patients with ovarian cancer divided into four equally sized strata. Each synthetic dataset was characterized by a different degree of strata separation *δ* (**Supplementary Fig. S1**). RedFuMOS achieved the highest AUROC across all scenarios (**Fig. 2**), demonstrating consistently superior stratification performance compared with all benchmarked state-of-the-art approaches. Notably, the improvement over competitors was most pronounced in scenarios with low to moderate inter-stratum separation. As expected, RedFuMOS performance improved progressively as the separation between simulated strata increased. Additionally, RedFuMOS exhibited more stable performance than CIMLR, IntNMF, PINSPlus, and moCluster, as evidenced by the larger standard deviations in AUROC observed for these competing approaches (**Fig. 2**).

**Fig. 2.**
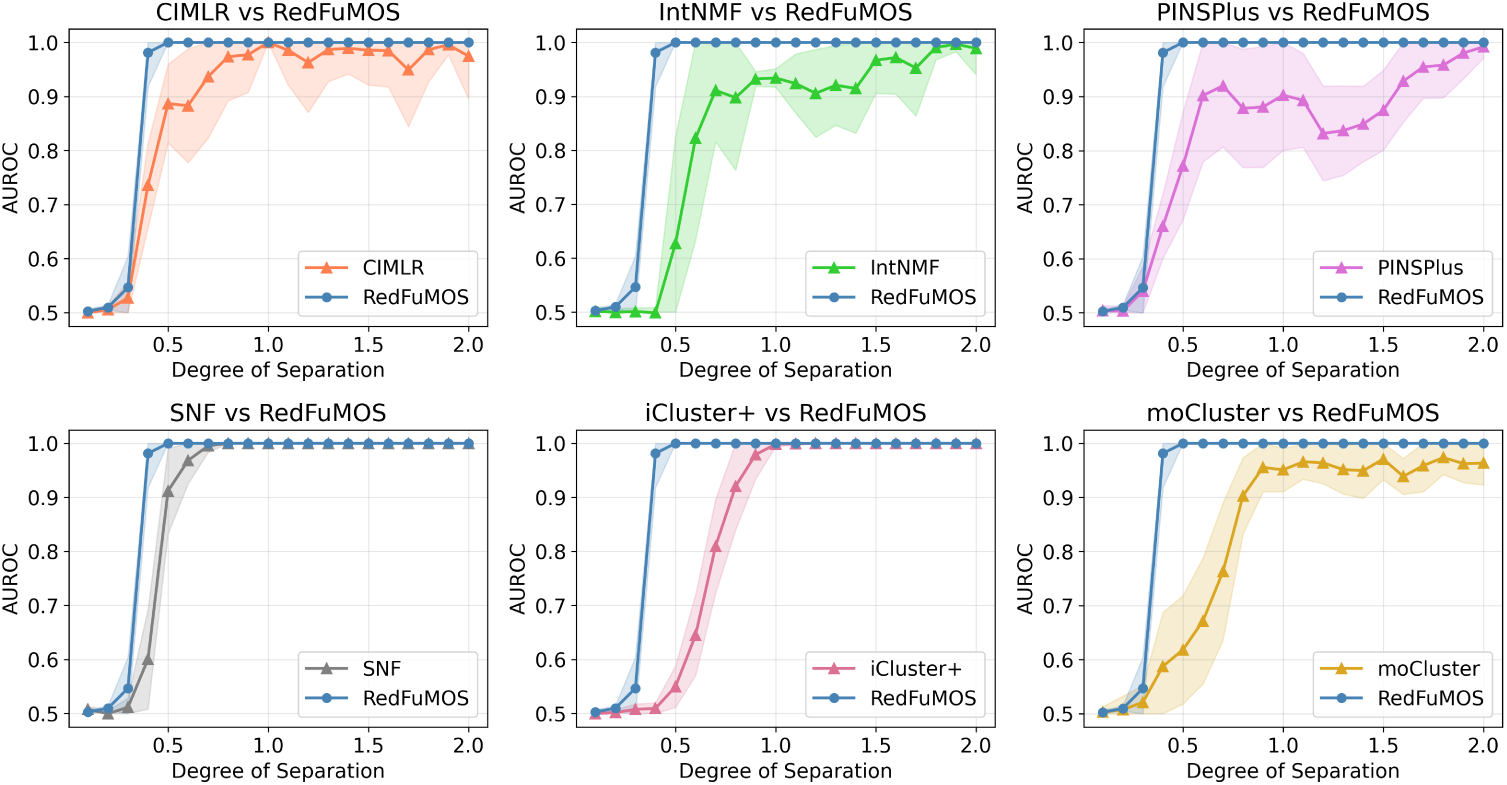
Benchmarking of multi*-omics* patient-stratification approaches. AUROC values obtained by RedFuMOS and six state-of-the-art approaches across increasing degrees of separation *δ* (x-axis) among simulated patient strata. AUROC values were calculated on synthetic datasets including 100 patients divided into four equally sized strata. The experiment was repeated 30 times, using different synthetic datasets. Points represent the mean AUROC values (y-axis) across the 30 repetitions, and shaded bands indicate *±* one standard deviation.

### Dimensionality reduction is crucial for patient stratification

In a second experiment, we assessed the scalability of RedFuMOS by simulating 10 datasets with a number of patients ranging from 100 to 1000, in increments of 100, with patients always divided into four equally sized strata. Running time scaled non-linearly with the number of patients (**Fig. 3A**), with the dimensionality reduction step accounting for the majority of the total running time, on average 81%. As expected, these observations are independent of the degree of strata separation (**Supplementary Fig. S2**).

**Fig. 3.**
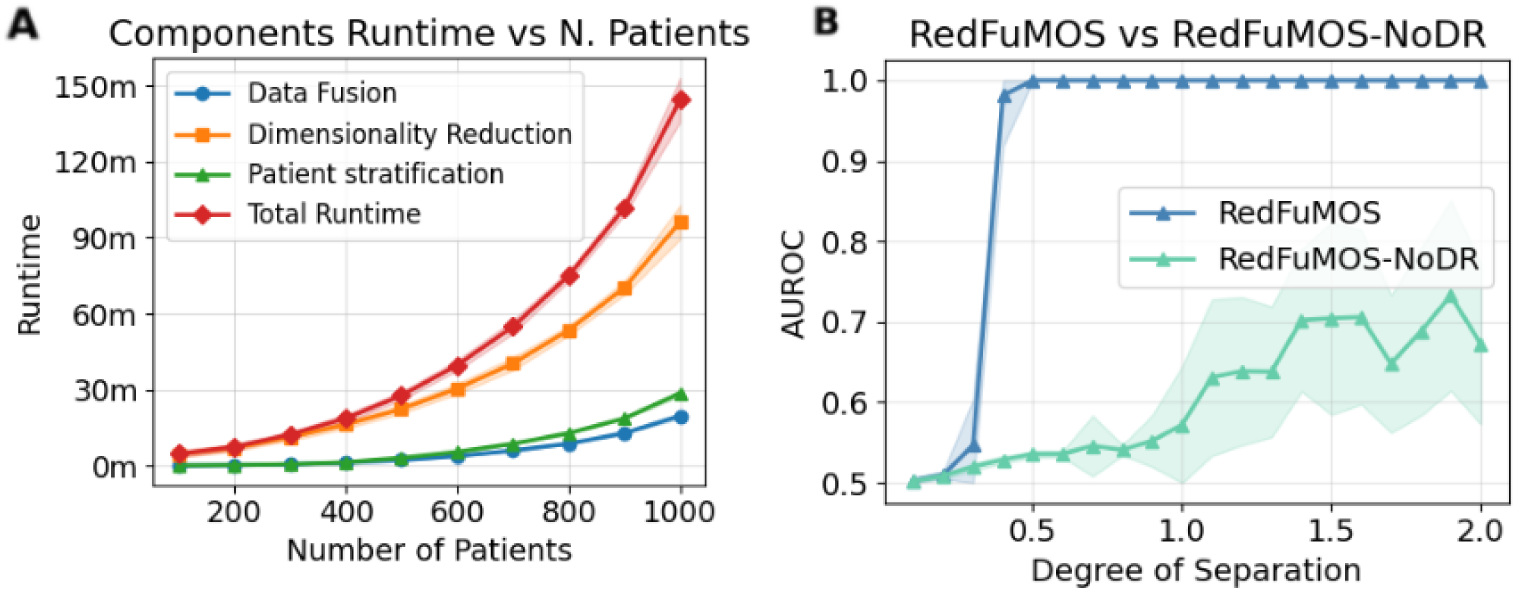
**(A) Scalability of RedFuMOS.** The total running time (red) and running times of each core component: data fusion (blue), dimensionality reduction (yellow), and patient stratification (green), as a function of sample size. Results are shown for synthetic datasets with a degree of separation of 0.5. For each sample size (x-axis), points represent the mean running time (y-axis) across the 10 repetitions, and shaded bands indicate *±* one standard deviation. **(B) Importance of dimensionality reduction.** Comparison between the complete RedFuMOS pipeline (blue) and a modified implementation in which the dimensionality reduction step was omitted (RedFuMOS-noDR, turquoise). For each degree of strata separation (x-axis), points represent the mean AUROC values (y-axis) across the 30 repetitions, and shaded bands indicate *±* one standard deviation.

Given the computational cost of dimensionality reduction, we investigated whether this step could be omitted without affecting stratification results. To this aim, we used the synthetic datasets generated for benchmarking to perform a third experiment, in which we compared the described version of RedFuMOS with a version where the dimensionality reduction step was omitted. We observed a substantial drop in performance (**Fig. 3B**), confirming that this step is essential for accurate stratification.

### RedFuMOS identifies clinically relevant strata in a real-world Ph+CML cohort

To evaluate the performance of RedFuMOS in a real-world setting, we used data from 40 patients with Ph+CML for whom two *-omics* layers were available: gut microbiome composition (*n*_genera_ = 86) and circulating cytokine levels (*n*_cytokines_ = 27). We selected these *-omics* layers because it has already been reported that treatment-related chronic inflammation increases intestinal permeability, with leaked microbial products stimulating cytokine production, which, in turn, intensifies intestinal mucosal damage [30]. Our objective was to understand whether RedFuMOS could separate patients who presented with a deep molecular response, a fundamental requisite for suspending therapy, from those who did not, and whether the obtained strata were associated with any biomarker signature.

RedFuMOS identified two patient strata including 27 and 10 patients, respectively, along with three patients labelled as outliers. (**Fig. 4 A**). The largest stratum included a larger proportion of patients with a deep molecular response (74%) than the smaller one (43%), corresponding to an odds ratio of 0.16 (95% confidence interval = 0.02-0.93, Fisher exact test P value = 0.02).

**Fig. 4.**
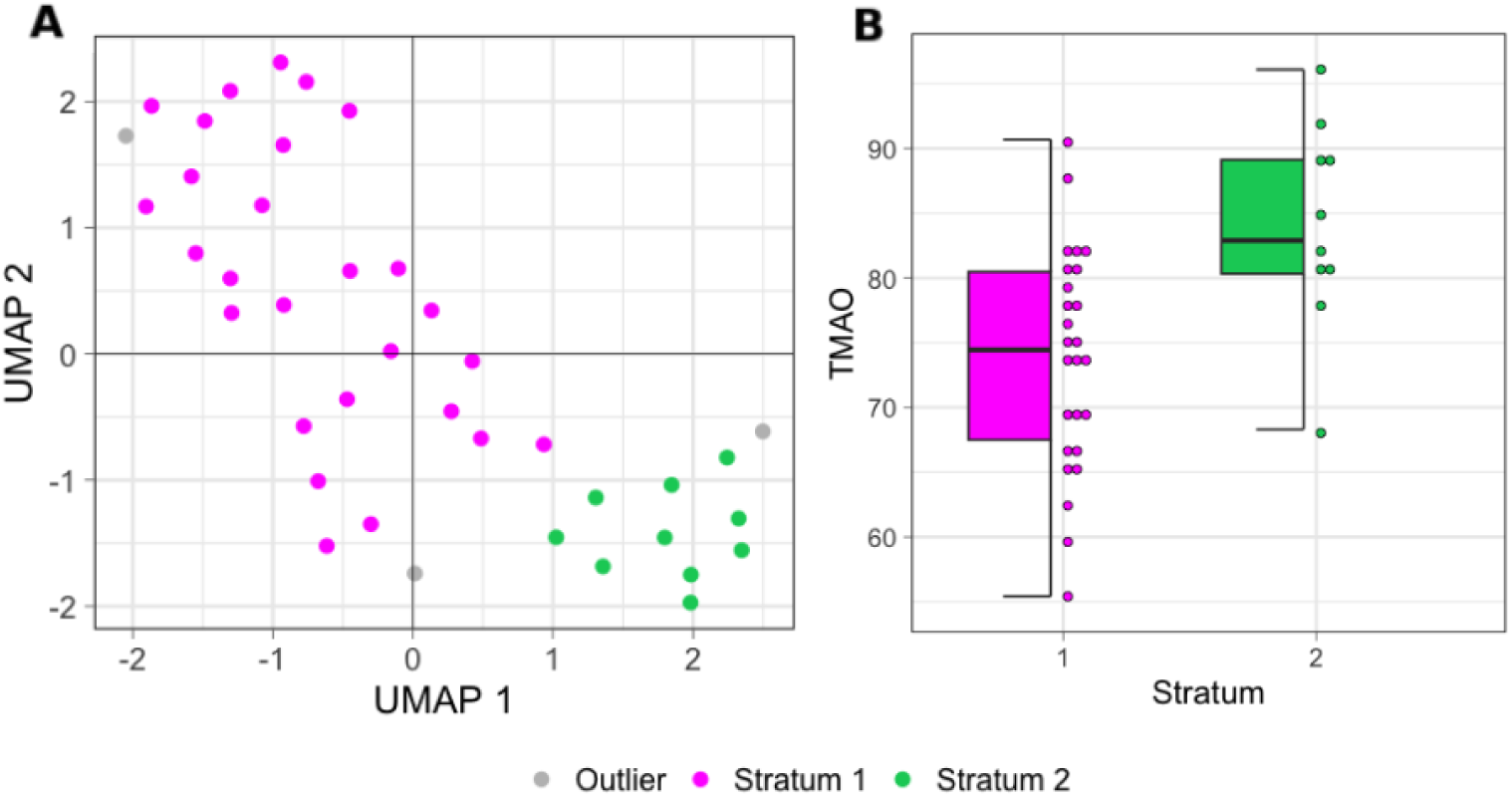
**(A) Stratification of patients with Ph+CML.** The dotplot shows the patient stratification results obtained by RedFuMOS on 40 patients with Ph+CML for whom two *-omics* layers were available. Patients are plotted according to their UMAP embedding, which was calculated during the dimensionality reduction step. **(B) Difference in TMAO levels between strata.** The boxplot shows the distribution of TMAO levels in the two strata identified by RedFuMOS.

To further characterize the two strata, we sought associations with demographic information (age and sex) and four plasma biomarkers (CRP, fibrinogen, DAO, and TMAO), identifying statistically significant differences in the levels of TMAO (Kruskal-Wallis P value = 0.004; **Fig. 4 B**). Additionally, we identified an association with age that did not survive multiple testing correction (Kruskal-Wallis P value = 0.01; **Supplementary Fig. S3**). These differences likely stem from the *-omics* layers we used in our experiment. Indeed, age is linked to modifications in the circulating pattern of pro- and anti-inflammatory cytokines [31], and TMAO is produced from the metabolism of dietary choline and L-carnitine by intestinal microbiota [32]. Interestingly, TMAO has been linked to cardiovascular risk [33], a common comorbidity in patients with Ph+CML [34], with previous observations also suggesting that plasma TMAO levels are positively correlated with age [35].

Additionally, we investigated whether similar results could have been generated by using a standard stratification approach on a single *-omics* layer. Thus, we ran HDBSCAN first on the microbiome layer, failing to identify any strata, and then on the cytokines layer, where we could identify two strata including four and 18 patients, respectively (18 patients were labelled as outliers). However, the identified strata were associated neither with a deep molecular response nor with any clinical variable or biomarker (P value *>* 0.1).

## Discussion

Identifying homogeneous patient strata is critical to enabling precision medicine. Since disease phenotypes arise from interaction of biological processes, patient stratification may benefit from the joint analysis of complementary multi*-omics* and clinical data.

In this study, we introduced RedFuMOS, a three-step approach for mixed data and multi-*omics* patient stratification based on similarity-network fusion, dimensionality reduction, and density-based clustering. Across all simulated scenarios, RedFuMOS outperformed six state-of-the-art approaches showing consistently superior stratification performance. Additionally, when used on a small cohort of patients with Ph+CML, RedFuMOS identified a stratum of younger patients with a higher likelihood of a deep molecular response and lower TMAO levels, who may have a reduced risk of developing cardiovascular disease following therapy [34], an insight that could not have been obtained using a single *-omics* layer alone.

A first distinctive feature of RedFuMOS is the inclusion of a dimensionality reduction step, which our experimentation showed to be critical for accurate patient stratification. Omitting this step resulted in a drop of performance, and we hypothesise that RedFuMOS outperformed well-performing approaches, such as iCluster+ and SNF (upon which RedFuMOS is built), thanks to its inclusion. High dimensional multi-*omics* data often contain noisy information that can hide the underlying patient structure. Dimensionality reduction approaches can mitigate this issue allowing a more compact representation while preserving key characteristics of the original data, such as data variance in PCA, or local structure in t-SNE and UMAP. This has been shown to improve downstream task, including clustering and their clinical characterization [36–38]. However, this step was also the most computationally demanding, highlighting a trade-off between stratification performance and computational efficiency.

A second feature is that RedFuMOS does not make any assumption on the data distribution, allowing for non-linear relationships, and thus outperforming linear-based approaches, such as IntNMF and MoCluster, across all benchmarked scenarios. This flexibility is crucial because relationship between *-omics* layers are often non-linear. For instance, correlation between mRNA expression levels and protein abundances are usually poor, due to post-transcriptional, translational and protein degradation regulation [39]. Additionally, environmental factors and feedback loops contribute to non-linearity, as highlighted by the study of gene-by-environment interactions [40]. Furthermore, some biological pathways often exhibit “switch-like” behaviour, where a change in a trait (*e.g.*, expression of a transcription factor-coding gene) does not influence a second one (*e.g.*, expression of the transcription factor-regulated gene) until a certain threshold is reached, after which a massive shift occurs [41]. This is particularly important, but not limited to, cancer and disease data, which have evolved a larger regulatory non-linearity than expected [42].

A third feature of RedFuMOS is that it does not require the number of patient strata to be specified *a priori*. This is particularly useful in exploratory studies, where the number of clinically relevant groups is rarely known in advance.

RedFuMOS can also handle multiple heterogeneous data layers allowing the use of layer-specific distance measure. This provides a common patient-similarity representation while retaining characteristics specific for each data type.

Finally, RedFuMOS incorporates k-NN imputation to address missing values, and applies an optimization procedure for the automated setting of hyperparameters by maximising the Silhouette score, minimizing the need for manual tuning.

Although we applied RedFuMOS to stratify cancer patients, it can also be applied to other patient populations, including healthy individuals or model organisms, for which multi-layer data are available.

RedFuMOS is available as an R package at http://github.com/delucasara/RedFuMOS. To facilitate future benchmarking efforts, all synthetic datasets are publicly available on Zenodo (https://zenodo.org/records/18451096).

This study has some limitations. First, the computational time increases significantly with sample size, because of dimensionality reduction step and hyperparameter optimization. This may limit application to very large cohorts. Nevertheless, for datasets comprising up to 1,000 patients, RedFuMOS completed the entire analysis in less than 2.5 hours on a standard laptop. Second, the benchmarking analysis was based on synthetic datasets containing only numerical molecular layers and therefore did not evaluate performance with mixed-type clinical variables. Third, the use of synthetic ovarian cancer data may not capture the full complexity of other diseases or data-generating processes. Finally, the real-world application included only 40 patients from a single centre and should be replicated in larger, independent cohorts. Although these analyses demonstrate RedFuMOS flexibility across several molecular data types, additional validation is needed for other modalities and for settings involving a larger number of layers.

## Conclusions

RedFuMOS provides a flexible framework for integrating heterogeneous multi*-omics* and clinical data to identify patient strata without requiring the number of strata to be specified *a priori* while modelling non-linear relationships. By accommodating mixed-type data, it addresses key limitations of existing methods, which often focus exclusively on molecular layers and overlook clinically relevant information. In our experimentation, RedFuMOS consistently outperformed several state-of-the-art integration methods and individual data layers, demonstrating that the combination of data fusion, dimensionality reduction, and density-based clustering yields robust and clinically meaningful patient strata These findings support the importance of using multi*-omics* and clinical data-based stratification approaches to improve patient stratification and support precision medicine.

## Abbreviations

AUROC: Area Under the Receiver Operating Characteristic
BCR-ABL1IS: Breakpoint Cluster Region - Abelson murine leukemia viral oncogene homolog 1 International Scale
CIMLR: Cancer Integration via Multi-kernel LeaRning
CML: Chronic Myeloid Leukaemia
CRP: C-Reactive Protein
DAO: Diamine Oxidase
HDBSCAN: Hierarchical Density-Based Spatial Clustering of Applications with Noise
IntNMF: Integrative Non-negative Matrix Factorization
k-NN: k-Nearest Neighbours
MR4: Major Molecular Response 4
PCA: Principal Component Analysis
Ph+CML: Philadelphia chromosome-positive Chronic Myeloid Leukaemia
RedFuMOS: Reduced Fusion of Multi-Omics Stratification
SNF: Similarity Network Fusion
t-SNE: t-distributed Stochastic Neighbor Embedding
TCGA: The Cancer Genome Atlas
TMAO: Trimethylamine N-Oxide
UMAP: Uniform Manifold Approximation and Projection

## Declarations

### Ethics approval and consent to participate

The study involving patients with Philadelphia positive chronic myeloid leukaemia was approved by the Territorial Ethics Committee of *A.O.U. Città della Salute e della Scienza di Torino* (protocol no. CE 209/2022, approval date 2022-05-02). The study protocol for patients with Philadelphia positive chronic myeloid leukaemia was registered at ClinicalTrials.gov (Identifier: NCT06724536). All patients provided written informed consent.

### Data availability

The synthetic datasets generated for the *in silico* validation are available on Zenodo (https://doi.org/10.5281/zenodo.18451096). Data on patients with a diagnosis of Philadelphia positive chronic myeloid leukaemia are available to *bona fide* researchers under managed access due to governance constraints, and can be requested from the corresponding author.

### Code availability

RedFuMOS is available as an R package and distributed *via* GitHub (http://github.com/delucasara/RedFuMOS) with a MIT license.

### Competing interests

The authors have no conflicts of interest to declare.

### Funding

This study was partly funded by a Pfizer Global Medical Grant to CF (MICROBIOLMC).

### Authors’ contributions

SDL, AV, and PB designed RedFuMOS. SDL implemented RedFuMOS and performed all computational experiments. AV designed the *in silico* validation. CF collected data on patients with a diagnosis of Philadelphia positive chronic myeloid leukaemia and contributed to the interpretation of the results. SDL and AV wrote the manuscript. All authors read, commented, and approved the final manuscript.

## Acknowledgements

We would like to thank Mr Sandro Gepiro Contaldo, who helped with the design of **Fig.1**.

## Notes

### Competing Interest Statement

The authors have declared no competing interest.

### Clinical Trial

NCT06724536

### Author Declarations

The study involving patients with Philadelphia positive chronic myeloid leukaemia was approved by the Territorial Ethics Committee of A.O.U. Citta' della Salute e della Scienza di Torino (protocol no. CE 209/2022, approval date 2022-05-02). The study protocol for patients with Philadelphia positive chronic myeloid leukaemia was registered at ClinicalTrials.gov (Identifier: NCT06724536). All patients provided written informed consent.

## References

[1] Menyhárt, O., Győrffy, B.: Multi-omics approaches in cancer research with applications in tumor subtyping, prognosis, and diagnosis. Computational and Structural Biotechnology Journal 19, 949–960 (2021) 10.1016/j.csbj.2021.01.009

[2] Wang, R.C., Wang, Z.: Precision Medicine: Disease Subtyping and Tailored Treatment. Cancers 15(15), 3837 (2023) 10.3390/cancers15153837

[3] Chen, C., Wang, J., Pan, D., Wang, X., Xu, Y., Yan, J., Wang, L., Yang, X., Yang, M., Liu, G.: Applications of multi-omics analysis in human diseases. MedComm 4(4), 315 (2023) 10.1002/mco2.315

[4] Tran, D., Nguyen, H., Pham, V.-D., Nguyen, P., Nguyen Luu, H., Minh Phan, L., Blair DeStefano, C., Jim Yeung, S.-C., Nguyen, T.: A comprehensive review of cancer survival prediction using multi-omics integration and clinical variables. Briefings in Bioinformatics 26(2), 150 (2025) 10.1093/bib/bbaf150

[5] Picard, M., Scott-Boyer, M.-P., Bodein, A., Périn, O., Droit, A.: Integration strategies of multi-omics data for machine learning analysis. Computational and Structural Biotechnology Journal 19, 3735–3746 (2021) 10.1016/j.csbj.2021.06.030

[6] Shen, R., Olshen, A.B., Ladanyi, M.: Integrative clustering of multiple genomic data types using a joint latent variable model with application to breast and lung cancer subtype analysis. Bioinformatics 25(22), 2906–2912 (2009) 10.1093/bioinformatics/btp543

[7] Mo, Q., Wang, S., Seshan, V.E., Olshen, A.B., Schultz, N., Sander, C., Powers, R.S., Ladanyi, M., Shen, R.: Pattern discovery and cancer gene identification in integrated cancer genomic data. Proceedings of the National Academy of Sciences 110(11), 4245–4250 (2013)

[8] Nguyen, H., Shrestha, S., Draghici, S., Nguyen, T.: PINSPlus: a tool for tumor subtype discovery in integrated genomic data. Bioinformatics 35(16), 2843–2846 (2019) 10.1093/bioinformatics/bty1049

[9] Chalise, P., Fridley, B.L.: Integrative clustering of multi-level ‘omic data based on non-negative matrix factorization algorithm. PLOS ONE 12(5), 1–18 (2017) 10.1371/journal.pone.0176278

[10] Meng, C., Helm, D., Frejno, M., Kuster, B.: moCluster: Identifying Joint Patterns Across Multiple Omics Data Sets. Journal of Proteome Research 15(3), 755–765 (2016) 10.1021/acs.jproteome.5b00824

[11] Ramazzotti, D., Lal, A., Wang, B., Batzoglou, S., Sidow, A.: Multi-omic tumor data reveal diversity of molecular mechanisms that correlate with survival. Nature Communications 9(1), 4453 (2018) 10.1038/s41467-018-06921-8

[12] Wang, B., Mezlini, A.M., Demir, F., Fiume, M., Tu, Z., Brudno, M., Haibe-Kains, B., Goldenberg, A.: Similarity network fusion for aggregating data types on a genomic scale. Nature Methods 11(3), 333–337 (2014) 10.1038/nmeth.2810

[13] Ye, X., Shi, T., Huang, D., Sakurai, T.: Multi-omics clustering by integrating clinical features from large language model. Methods 239, 64–71 (2025) 10.1016/j.ymeth.2025.03.017

[14] Siam, M.B.H., Khan, M.R., Elahe, M.F., Arman, M.S., Akter, S.: Effects of similarity networks in graph-based multi-omics classification. Plos one 21(3), 0344754 (2026)

[15] Liu, P., Yuan, H., Ning, Y., Chakraborty, B., Liu, N., Peres, M.A.: A modified and weighted Gower distance-based clustering analysis for mixed type data: a simulation and empirical analyses. BMC Medical Research Methodology 24(1), 305 (2024) 10.1186/s12874-024-02427-8

[16] McKnight, D.T., Huerlimann, R., Bower, D.S., Schwarzkopf, L., Alford, R.A., Zenger, K.R.: Methods for normalizing microbiome data: An ecological perspective. Methods in Ecology and Evolution 10(3), 389–400 (2019) 10.1111/2041-210X.13115

[17] Jin, X., Jiang, Q., Chen, Y., Lee, S.-J., Nie, R., Yao, S., Zhou, D., He, K.: Similarity/dissimilarity calculation methods of DNA sequences: A survey. Journal of Molecular Graphics and Modelling 76, 342–355 (2017) 10.1016/j.jmgm.2017.07.019

[18] Jaskowiak, P.A., Campello, R.J., Costa, I.G.: On the selection of appropriate distances for gene expression data clustering. BMC Bioinformatics 15(2), 2 (2014) 10.1186/1471-2105-15-S2-S2

[19] Nguyen, V.-A., Liò, P.: Measuring similarity between gene expression profiles: a Bayesian approach. BMC Genomics 10(3), 14 (2009) 10.1186/1471-2164-10-S3-S14

[20] Qi, Z., Voit, E.O.: Strategies for Comparing Metabolic Profiles: Implications for the Inference of Biochemical Mechanisms from Metabolomics Data. IEEE/ACM Transactions on Computational Biology and Bioinformatics 14(6), 1434–1445 (2017) 10.1109/TCBB.2016.2586065

[21] Hotelling, H.: Analysis of a complex of statistical variables into principal components. Journal of educational psychology 24(6), 417 (1933)

[22] Maaten, L., Hinton, G.: Visualizing data using t-sne. Journal of Machine Learning Research 9(86), 2579–2605 (2008)

[23] Hinton, G.E., Roweis, S.: Stochastic neighbor embedding. Advances in neural information processing systems 15 (2002)

[24] McInnes, L., Healy, J., Melville, J.: Umap: Uniform manifold approximation and projection for dimension reduction. arXiv preprint arXiv:1802.03426 (2018)

[25] McInnes, L., Healy, J., Astels, S.: HDBSCAN: Hierarchical density based clustering. Journal of Open Source Software 2(11), 205 (2017) 10.21105/joss.00205

[26] Rousseeuw, P.J.: Silhouettes: A graphical aid to the interpretation and validation of cluster analysis. Journal of Computational and Applied Mathematics 20, 53–65 (1987) 10.1016/0377-0427(87)90125-7

[27] Thrun, M.C., Märte, J., Stier, Q.: Analyzing Quality Measurements for Dimensionality Reduction. Machine Learning and Knowledge Extraction 5(3), 1076– 1118 (2023) 10.3390/make5030056

[28] Chalise, P., Raghavan, R., Fridley, B.L.: InterSIM: Simulation tool for multiple integrative ‘omic datasets’. Computer Methods and Programs in Biomedicine 128, 69–74 (2016) 10.1016/j.cmpb.2016.02.011

[29] Network, T.C.G.A.R.: Integrated genomic analyses of ovarian carcinoma. Nature 474(7353), 609–615 (2011) 10.1038/nature10166

[30] Jalalifar, S., Bajelan, B., Mohammadi, R., Ghafoury, R., Kalhori, Z., Pooshang-Bagheri, K., Nekouian, R., Faranoush, M.: The impact of gut microbiota on leukemia and prospects for novel therapies. Infectious Medicine 5(1), 100239 (2026) 10.1016/j.imj.2026.100239

[31] Rea, I.M., Gibson, D.S., McGilligan, V., McNerlan, S.E., Alexander, H.D., Ross, O.A.: Age and age-related diseases: role of inflammation triggers and cytokines. Frontiers in immunology 9, 586 (2018)

[32] Zhou, Y., Zhang, Y., Jin, S., Lv, J., Li, M., Feng, N.: The gut microbiota derived metabolite trimethylamine N-oxide: its important role in cancer and other diseases. Biomedicine & Pharmacotherapy 177, 117031 (2024)

[33] Dean, Y.E., Rouzan, S.S., Pintado, J.J.L., Talat, N.E., Mohamed, A.R., Verma, S., Kamdi, Z.A., Gir, D., Helmy, A., Helmy, Z., et al.: Serum trimethylamine N-oxide levels among coronary artery disease and acute coronary syndrome patients: a systematic review and meta-analysis. Annals of Medicine and Surgery 85(12), 6123–6133 (2023)

[34] Santoro, M., Mancuso, S., Accurso, V., Di Lisi, D., Novo, G., Siragusa, S.: Cardiovascular issues in tyrosine kinase inhibitors treatments for chronic myeloid leukemia: a review. Frontiers in Physiology 12, 675811 (2021)

[35] Wang, Z., Levison, B.S., Hazen, J.E., Donahue, L., Li, X.-M., Hazen, S.L.: Measurement of trimethylamine-N-oxide by stable isotope dilution liquid chromatography tandem mass spectrometry. Analytical biochemistry 455, 35–40 (2014)

[36] Wani, A.A.: Comprehensive review of dimensionality reduction algorithms: challenges, limitations, and innovative solutions. PeerJ Computer Science 11, 3025 (2025)

[37] Sun, S., Zhu, J., Ma, Y., Zhou, X.: Accuracy, robustness and scalability of dimensionality reduction methods for single-cell RNA-seq analysis. Genome biology 20(1), 269 (2019)

[38] Cantini, L., Zakeri, P., Hernandez, C., Naldi, A., Thieffry, D., Remy, E., Baudot, A.: Benchmarking joint multi-omics dimensionality reduction approaches for the study of cancer. Nature communications 12(1), 124 (2021)

[39] Vogel, C., Marcotte, E.M.: Insights into the regulation of protein abundance from proteomic and transcriptomic analyses. Nature reviews genetics 13(4), 227–232 (2012)

[40] Ritz, B.R., Chatterjee, N., Garcia-Closas, M., Gauderman, W.J., Pierce, B.L., Kraft, P., Tanner, C.M., Mechanic, L.E., McAllister, K.: Lessons learned from past gene-environment interaction successes. American Journal of Epidemiology 186(7), 778–786 (2017) 10.1093/aje/kwx230

[41] Siegal-Gaskins, D., Mejia-Guerra, M.K., Smith, G.D., Grotewold, E.: Emergence of switch-like behavior in a large family of simple biochemical networks. PLoS Computational Biology 7(5), 1002039 (2011) 10.1371/journal.pcbi.1002039

[42] Manicka, S., Johnson, K., Levin, M., Murrugarra, D.: The nonlinearity of regulation in biological networks. NPJ Systems Biology and Applications 9(1), 10 (2023)

