## Supplementary Material for "RedFuMOS: A novel approach for multi*-omics* and clinical data-driven patient stratification"

### Supplementary Figures

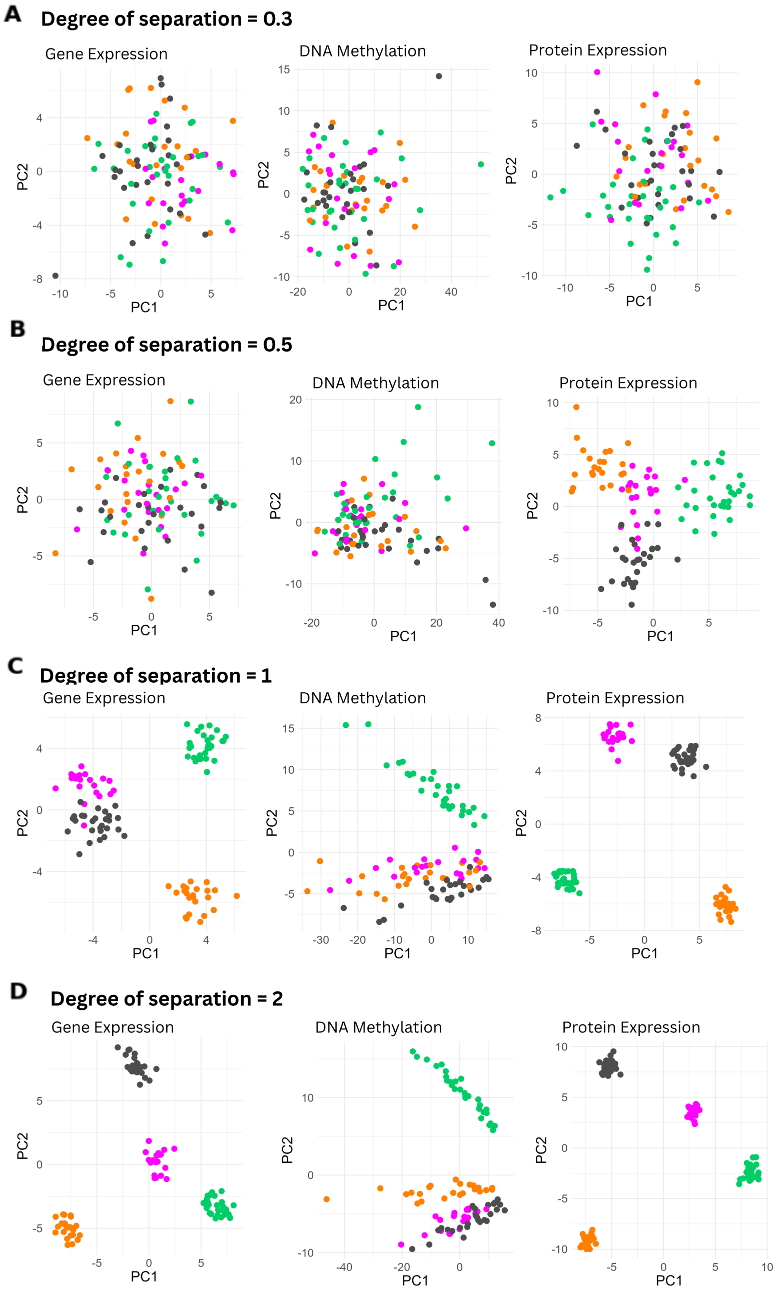


**Supplementary Figure S1**: **Examples of synthetic datasets**. The scatterplots show the first two principal components (PCs) for the *-omics* layers generated by the *InterSIM* R package [doi:10.1016/j.cmpb.2016.02.011], namely gene expression levels (left), DNA methylation (middle), and protein expression levels (right). Each panel shows a different degree of separation $\delta$ between strata, as indicated in the panel title. Higher degrees of separation correspond to greater separation between strata. Each dataset included 100 patients with ovarian cancer, divided into four equally sized strata (in magenta, green, grey, and orange).

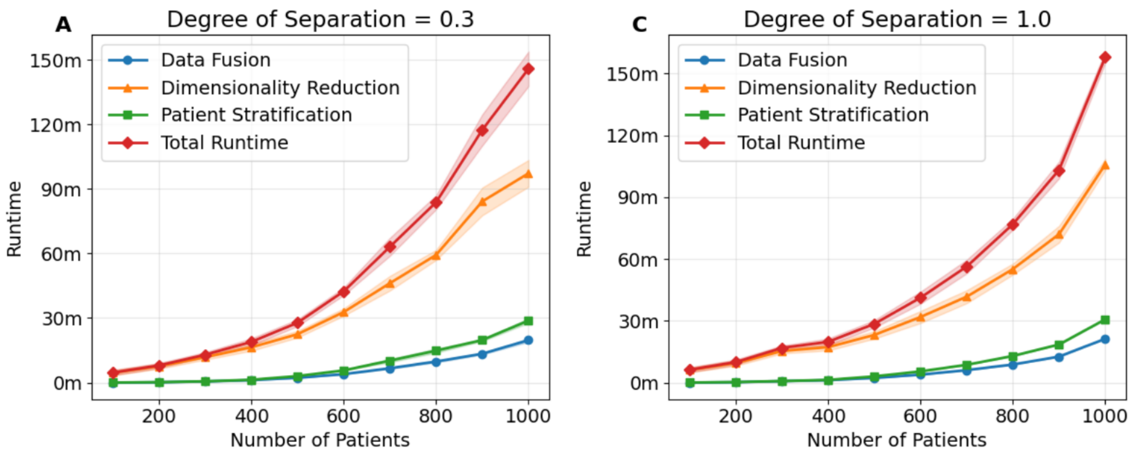


**Supplementary Figure S2**: **Scalability of RedFuMOS.** The total running time (red) and running times of each core component: data fusion (blue), dimensionality reduction (yellow), and patient stratification (green), as a function of sample size. Results are shown for synthetic datasets with a degree of separation of 0.3 (Panel A, left) and 1 (Panel B, right). For each sample size (x-axis), points represent the mean running time (y-axis) across the 10 repetitions, and shaded bands indicate $\pm$ one standard deviation.

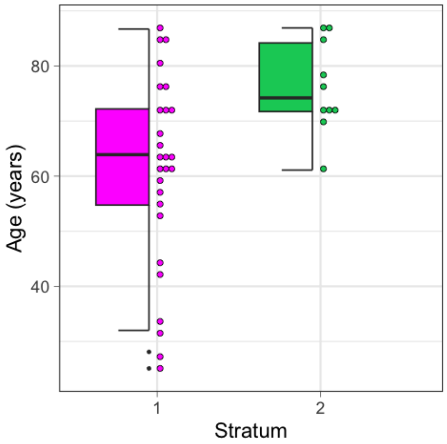


**Supplementary Figure S3**: **Difference in age between strata.** The box plot shows the distribution of age in the strata identified by RedFuMOS in the cohort of 40 patients with a diagnosis of Philadelphia-positive chronic myeloid leukaemia.

### Supplementary Tables

**Supplementary Table S1: State-of-the-art tools used for benchmarking.** The table lists each tool's name, its clickable DOI reference, the package version used, and a clickable link to the code repository from which it was downloaded.

| Name | DOI | Version | Downloaded from |
|---|---|---|---|
| CIMLR | 10.1038/s41467-018-06921-8 | 1.0.0 | GitHub |
| IntNMF | 10.1371/journal.pone.0176278 | 1.3.0 | CRAN |
| PINSPlus | 10.1093/bioinformatics/bty1049 | 2.0.7 | CRAN |
| SNFtool | doi.org/10.1038/nmeth.2810 | 2.3.1 | CRAN |
| iClusterPlus | 10.1093/biostatistics/kxx017 | 1.42.0 | Bioconductor |
| moCluster* | 10.1021/acs.jproteome.5b00824 | 1.40.0 | Bioconductor |

*Implemented in the `mogsa` R package
